# Mental health and substance use associated with hospitalization among people with laboratory confirmed diagnosis of COVID-19 in British Columbia: a population-based cohort study

**DOI:** 10.1101/2021.08.26.21262697

**Authors:** Héctor Alexander Velásquez García, James Wilton, Kate Smolina, Mei Chong, Drona Rasali, Michael Otterstatter, Caren Rose, Natalie Prystajecky, Samara David, Eleni Galanis, Geoffrey McKee, Mel Krajden, Naveed Zafar Janjua

## Abstract

**Background:** This study identified factors associated with hospital admission among people with laboratory-diagnosed COVID-19 cases in British Columbia.

**Methods:** This study was performed using the BC COVID-19 Cohort, which integrates data on all COVID-19 cases, hospitalizations, medical visits, emergency room visits, prescription drugs, chronic conditions and deaths. The analysis included all laboratory-diagnosed COVID-19 cases in British Columbia as of January 15^th^, 2021. We evaluated factors associated with hospital admission using multivariable Poisson regression analysis with robust error variance.

**Findings:** From 56,874 COVID-19 cases included in the analyses, 2,298 were hospitalized. Models showed significant association of the following factors with increased hospitalization risk: male sex (adjusted risk ratio (aRR)=1.27; 95%CI=1.17-1.37), older age (p-trend <0.0001 across age groups with a graded increase in hospitalization risk with increasing age [aRR 30-39 years=3.06; 95%CI=2.32-4.03, to aRR 80+years=43.68; 95%CI=33.41-57.10 compared to 20-29 years-old]), asthma (aRR=1.15; 95%CI=1.04-1.26), cancer (aRR=1.19; 95%CI=1.09-1.29), chronic kidney disease (aRR=1.32; 95%CI=1.19-1.47), diabetes (treated without insulin aRR=1.13; 95%CI=1.03-1.25, requiring insulin aRR=5.05; 95%CI=4.43-5.76), hypertension (aRR=1.19; 95%CI=1.08-1.31), injection drug use (aRR=2.51; 95%CI=2.14-2.95), intellectual and developmental disabilities (aRR=1.67; 95%CI=1.05-2.66), problematic alcohol use (aRR=1.63; 95%CI=1.43-1.85), immunosuppression (aRR=1.29; 95%CI=1.09-1.53), and schizophrenia and psychotic disorders (aRR=1.49; 95%CI=1.23-1.82). Among women of reproductive age, in addition to age and comorbidities, pregnancy (aRR=2.69; 95%CI=1.42-5.07) was associated with increased risk of hospital admission.

**Interpretation:** Older age, male sex, substance use, intellectual and developmental disability, chronic comorbidities, and pregnancy increase the risk of COVID-19-related hospitalization.

**Funding:** BC Centre for Disease Control, Canadian Institutes of Health Research.

**Research in context:** *Evidence before this study:* Factors such as older age, social inequities and chronic health conditions have been associated to severe COVID-19 illness. Most of the evidence comes from studies that don’t include all COVID-19 diagnoses in a jurisdiction), focusing on in-hospital mortality. In addition, mental illness and substance use were not evaluated in these studies. This study assessed factors associated with hospital admission among people with laboratory-diagnosed COVID-19 cases in British Columbia.

*Added value of this study:* In this population-based cohort study that included 56,874 laboratory-confirmed COVID-19 cases, older age, male sex, injection drug use, problematic alcohol use, intellectual and developmental disability, schizophrenia and psychotic disorders, chronic comorbidities and pregnancy were associated with the risk of hospitalization. Insulin-dependent diabetes was associated with higher risk of hospitalization, especially in the subpopulation younger than 40 years. To the best of our knowledge this is the first study reporting this finding, (insulin use and increased risk of COVID-19-related death has been described previously).

*Implications of all the available evidence:* Prioritization of vaccination in population groups with the above mentioned risk factors could reduce COVID-19 serious outcomes. The findings indicate the presence of the syndemic of substance use, mental illness and COVID-19, which deserve special public health considerations.

## Introduction

COVID-19 caused by SARS-CoV-2, affecting millions of people globally, resulted in a spectrum of health outcomes among those affected. Presentations range from asymptomatic and mild illnesses to severe disease that requires hospitalization with varying needs for more intensive levels of care.^1–3^

Elevated levels of hospitalization and need for healthcare surge capacity, particularly intensive care unit (ICU), have been key drivers of pandemic response planning and contributed to societal disruption.^3^ Studies have identified various demographic factors (e.g. being older in age, male) and chronic comorbidities (e.g. diabetes, cardiovascular disease (CVD), hypertension) as risk factors for hospitalization and other severe outcomes;^2^ however, most studies were conducted on patients presenting at hospitals. There are very few population-based studies investigating risk factors in the entire population of COVID-19 diagnosed individuals in a jurisdiction. This can lead to potential selection bias when characterizing risk factors. Older age has been identified as the strongest risk factor for severe disease along with various comorbidities,^4^ however, very few studies investigated relationship of substance use, intellectual disabilities and role of insulin dependent diabetes with the risk of severe outcomes.^5,6^

In addition, the clinical threshold to admit to hospital may vary across countries, especially early in the pandemic; hence, knowledge of factors associated with hospitalization in the Canadian context is important. Population-level hospitalization risk factor studies are also particularly essential for prioritization of interventions to reduce health system burden and maintain hospital capacity. In this study, we identified factors associated with hospital admission among people with COVID-19 infection in British Columbia (BC).

## Methods

### Study population

The study used data from the BC COVID-19 Cohort (BCC19C), which integrates data on all individuals tested for COVID-19 in BC, including all lab-diagnosed cases, COVID-19 hospital and ICU census, medical visits, other hospitalizations, emergency room visits, prescription drugs, chronic conditions and mortality (Supplementary data, Appendix A, Table S1). That BCC19C was established as a public health surveillance system under the BCCDC’s public health mandate. This study was reviewed and approved by the Behavioral Research Ethics Board at the University of British Columbia (approval # H20-02097).

The study population for this analysis included individuals who tested positive for SARS-CoV-2 by real-time reverse transcription–polymerase chain reaction (RT-PCR), from January 26^th^, 2020 to January 15^th^, 2021. The outcome of interest was hospitalization as the measure of case severity related to COVID-19 infection, defined as hospital admission in a BC acute care facility within 14 days after a positive SARS-CoV-2 test^7–9^. Patients residing in long term care facilities were excluded from the analyses as their transfer to hospitals was variable over time and across local regions. For women of reproductive age (15-49 years), hospital admissions related to COVID-19 were considered as such only if no obstetric-related hospitalization code was found in DAD, within 14 days of hospital admission (Appendix D).

#### Comorbidities

We examined the following chronic conditions: Alzheimer/dementia, asthma, chronic heart disease (CHD: acute myocardial infarct, angina, heart failure, ischemic myocardial infarct), chronic obstructive pulmonary disease (COPD), cirrhosis, chronic kidney disease (CKD), depression, diabetes (categorized as no-diabetes, treated without insulin, and requiring insulin), epilepsy, gout, hypertension, stroke (ischemic, haemorrhagic, transitory ischemic attack), mood and anxiety disorders, osteoarthritis, osteoporosis, parkinsonism, rheumatoid arthritis, substance use disorder, injection drug use (IDU), problematic alcohol use, cancer, immunosuppression, intellectual and developmental disabilities (IDD), and schizophrenia and psychotic disorders (SZP). Variable definitions and diagnostic codes are detailed in Supplementary data (Appendix C).

### Statistical analysis

We described the baseline characteristics of participants including age, sex, and pregnancy status.

We evaluated risk factors associated with hospital admission, calculating risk ratios through multivariable Poisson regression models with robust error variance.^10^ For age, analyses were conducted as continuous as well as categorized into groups. To assess population differences across time, the cohort was stratified according to two waves or time periods: January 26^th^ to August 1^st^, 2020, and August 2^nd^, 2020 to January 15^th^, 2021. Sensitivity analyses were performed by 1) stratifying the population by age group, and 2) by restricting the outcome to hospitalizations lasting more than two days to address severity. All statistical analyses were performed using R version 4.0.2.

## Results

The analysis included 56,874 COVID-19 cases diagnosed up to January 15^th^, 2021; 2,298 (4.0%) of them were admitted to the hospital. A slightly higher proportion of people diagnosed as COVID-19 positive were male (51.2%), with much higher proportion among people requiring hospital admission (58.5% vs. 50.9) (Table 1).The median age of COVID-19 cases was 35 years (IQR:24-50), while that of people requiring hospital admission was nearly twice that (66 years; IQR:53-78). The proportion of hospitalized cases increased with each 10 year age-groups increment, from 0.2% in the youngest subpopulation (<20 years) to 34.2% in the eldest group (80+ years) (RR=66.96; 95%CI=52.35-85.65). Pregnant women were more likely to be hospitalized than women who were not (2.5% vs. 1.1%).

**Table 1.**
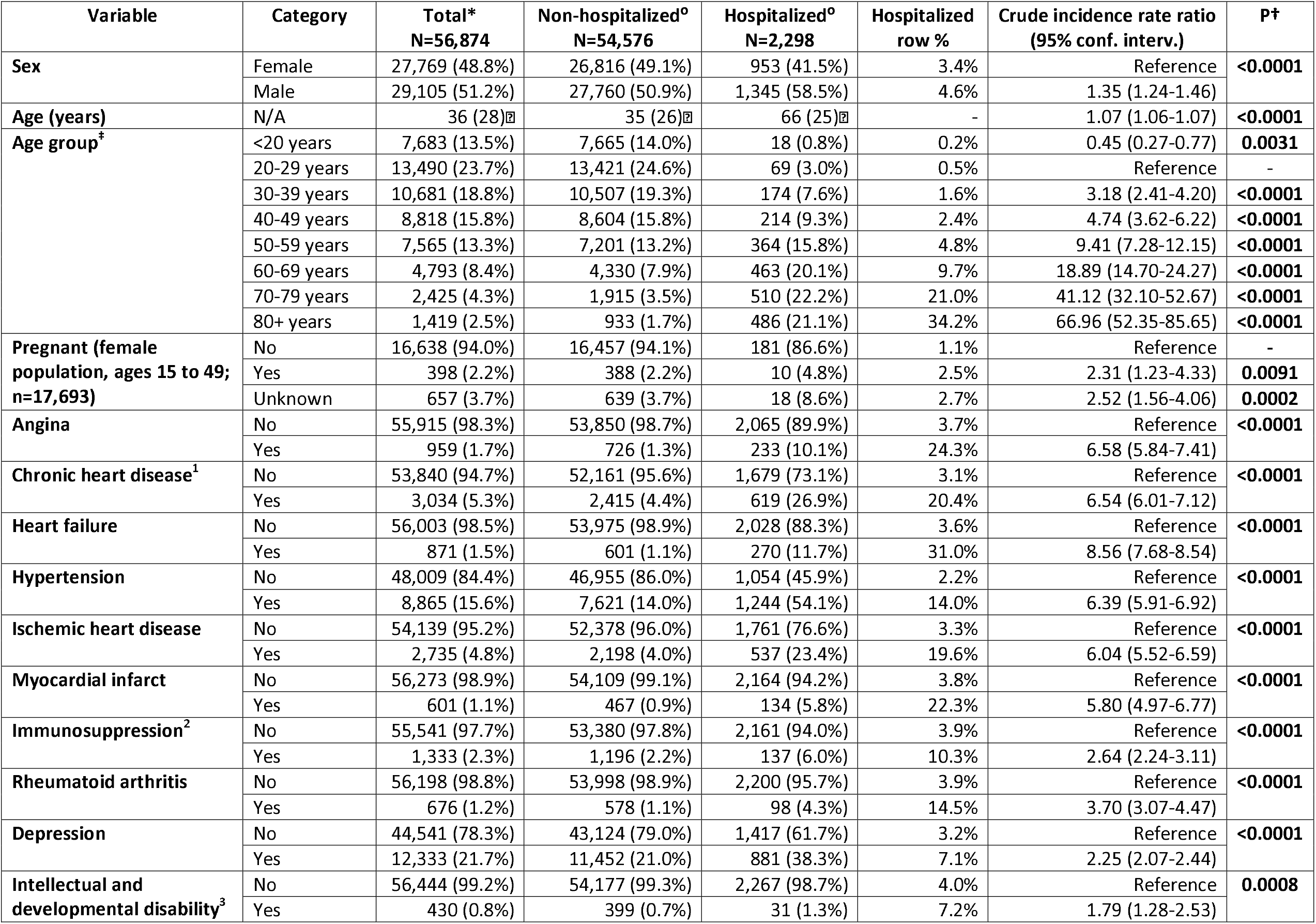
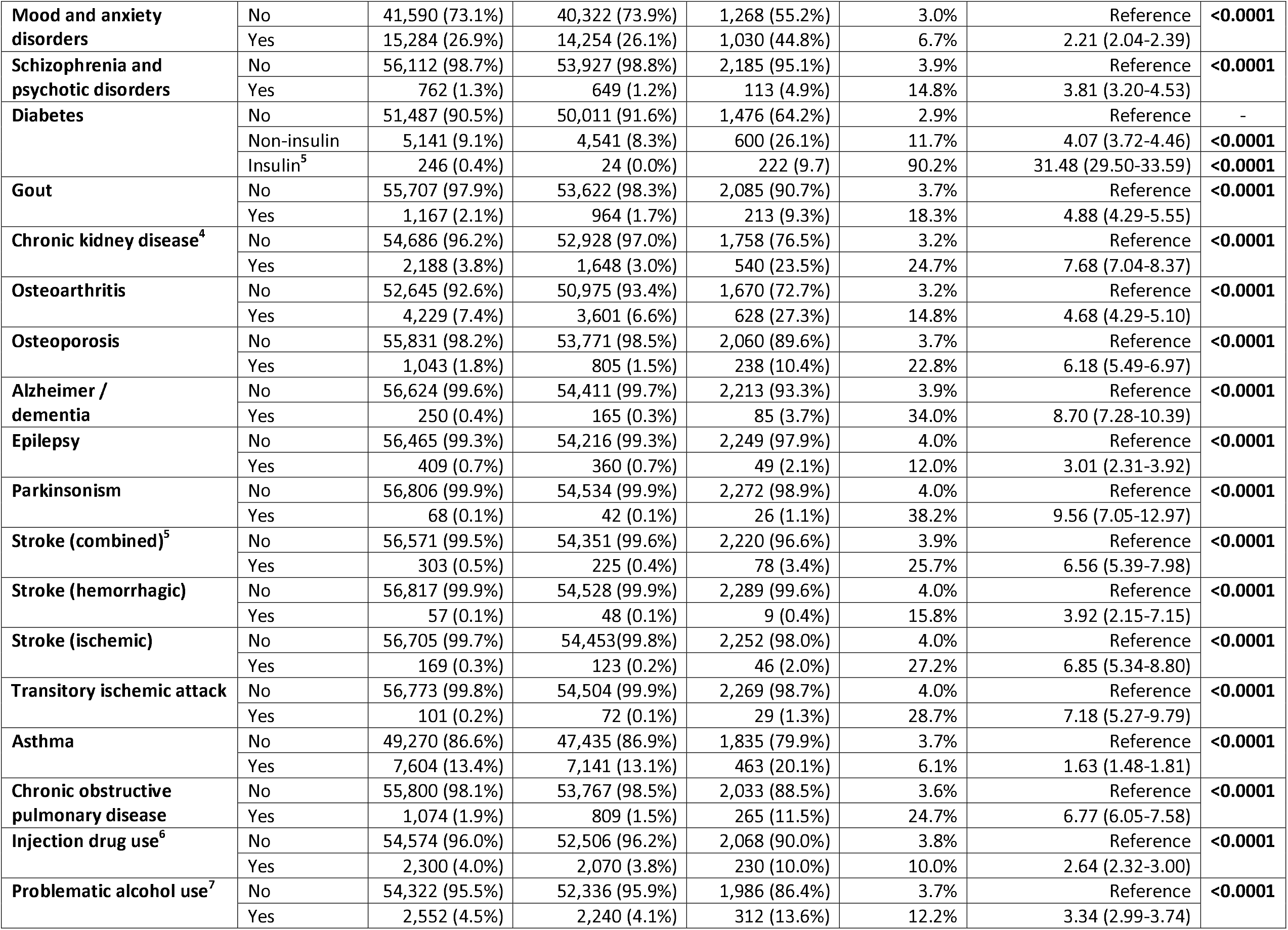
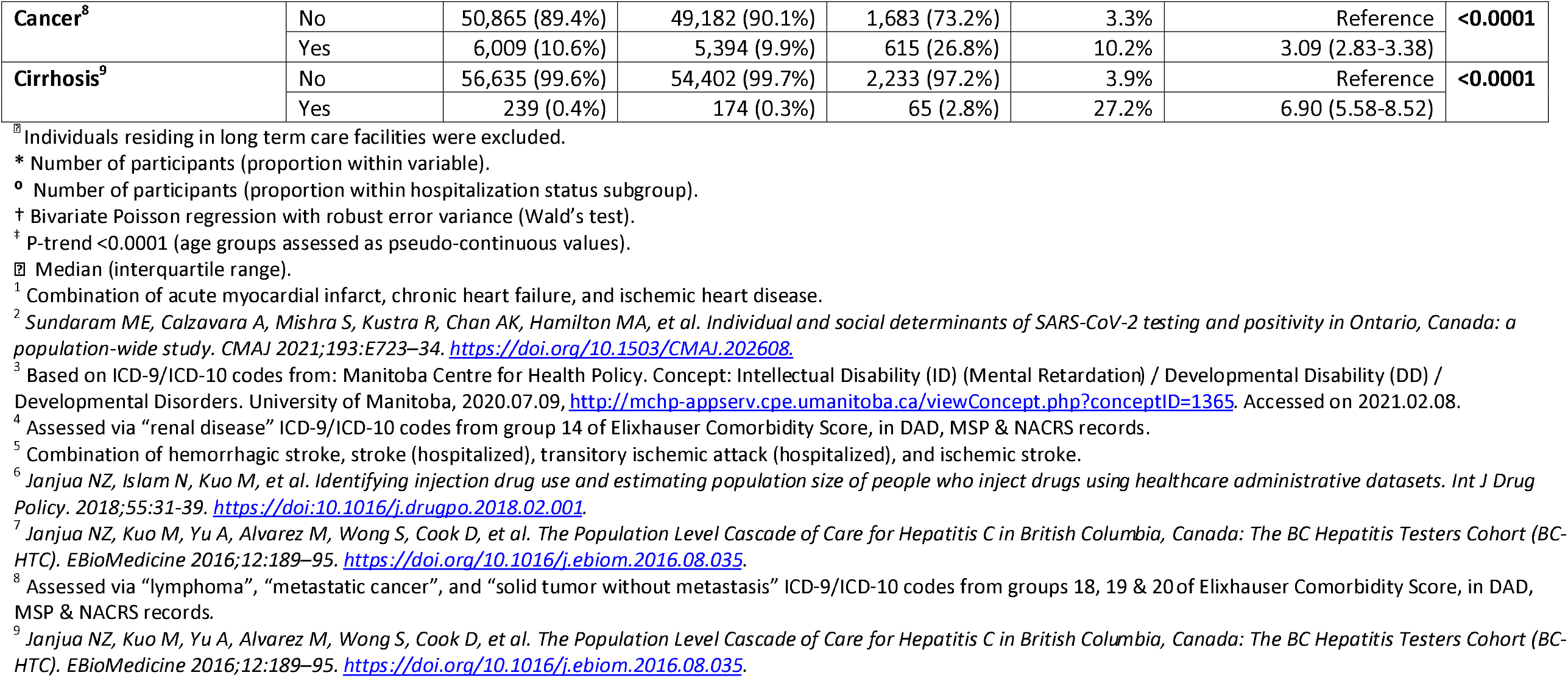
Distribution of characteristics in the BC COVID-19 cohort (confirmed cases, N= 56,874)^⍰^, according to hospitalization status.

### Chronic comorbidities as risk factors

The proportion of comorbidities among hospitalized individuals was higher than in those who did not require hospitalization (Table 1), including hypertension (54.1 vs. 14%), depression (38.3 vs. 21.7%), diabetes (35.8 vs. 8.3%), osteoarthritis (27.3 vs. 6.6%), CHD (26.9 vs. 4.4%), cancer (26.8 vs. 9.9%), CKD (23.5 vs. 3.0%), asthma (20.1 vs. 13.1%), substance use disorder (13.7 vs. 4.3%), problematic alcohol use (13.6 vs. 4.1%), heart failure (11.7 vs. 1.1%), COPD (11.5 vs. 1.5%), IDU (10 vs. 3.8%), and immunosuppression (6.0 vs. 2.2%).

In the adjusted multivariable Poisson regression model (Table 2; Figure 1), older age (p-trend <0.0001 across age groups with a graded increase in hospitalization risk with increasing age [aRR 30-39 years=3.06; 95%CI=2.32-4.03, to aRR 80+years=43.68; 95%CI=33.41-57.10 compared to 20-29 years-old]), male sex (aRR=1.27; 95%CI=1.17-1.37), asthma (aRR=1.15; 95%CI=1.04-1.26), cancer (1.19; 95%CI=1.09-1.29), CKD (RR=1.32; 95%CI=1.19-1.47), diabetes (treated without insulin aRR=1.13; 95%CI=1.03-1.25, requiring insulin aRR=5.05; 95%CI=4.43-5.76), hypertension (aRR=1.19; 95%CI=1.08-1.31), immunosuppression(1.30; 95%CI=1.10-1.54), IDU (aRR=2.51; 95%CI=2.14-2.95), IDD (aRR=1.67; 95%CI=1.05-2.66), problematic alcohol use (aRR=1.63; 95%CI=1.43-1.85), and SZP (aRR=1.49; 95%CI=1.23-1.82), were associated with increased hospitalization risk. The analysis by time period (Supplemental Tables S2, S3) did not show any remarkable differences in risk factors.

**Table 2.**
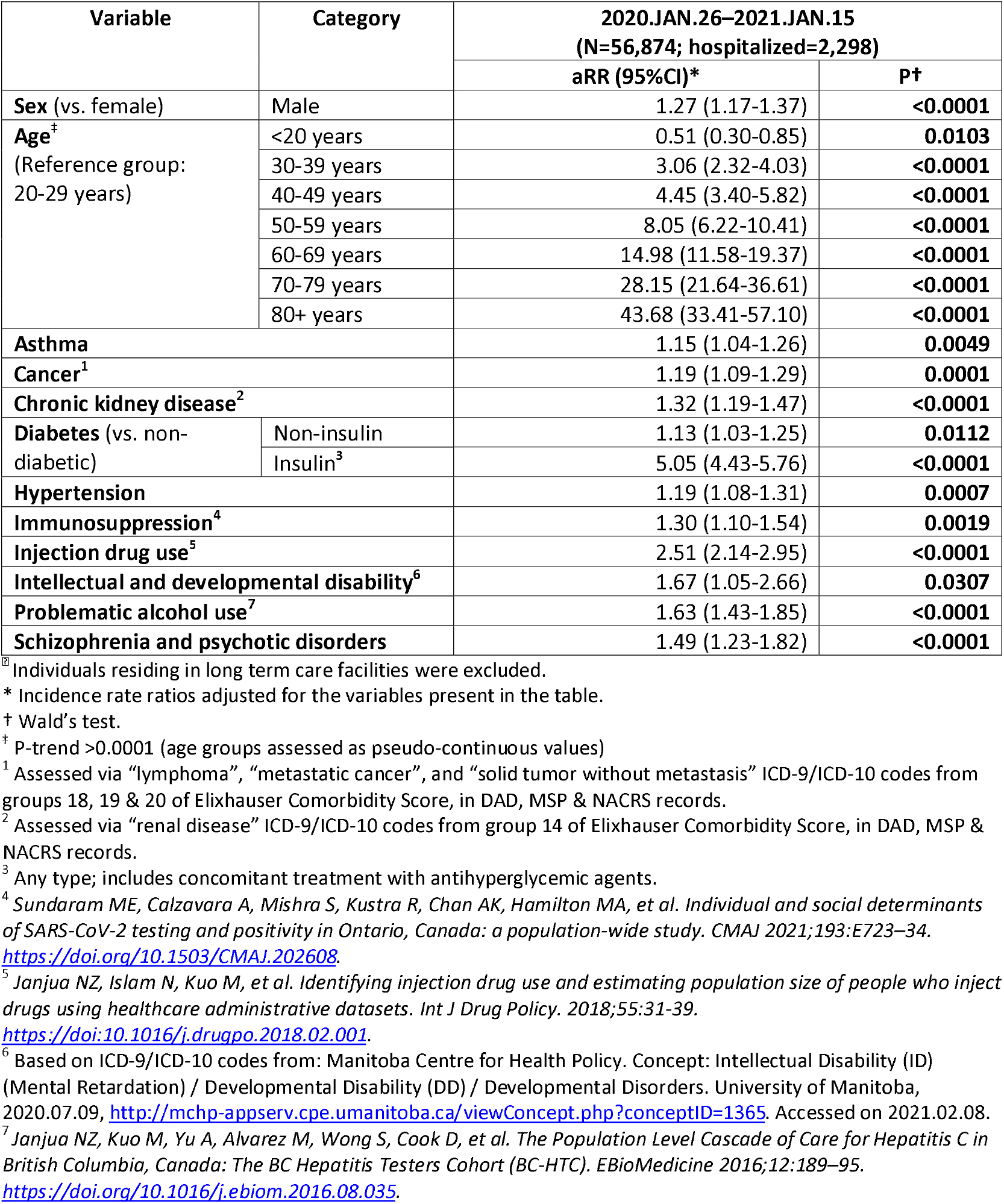
Factors associated with hospitalization status in multivariable Poisson regression analysis with robust error variance among confirmed cases, BC COVID-19 Cohort^⍰^.

**Figure 1.**
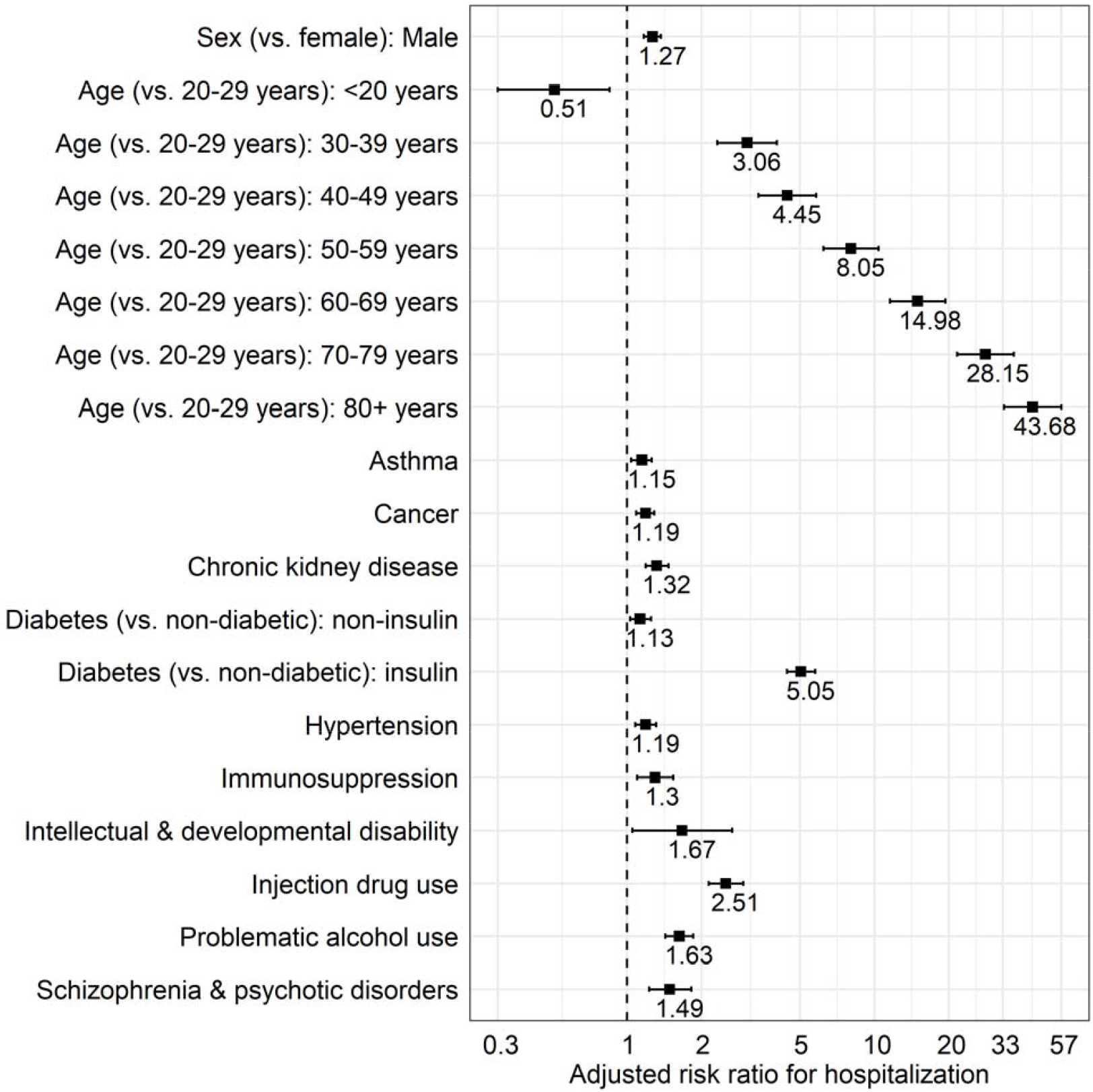
Multivariable model for factors associated with COVID-19-related hospitalization in British Columbia.

Among women of reproductive age(Table 3), pregnancy (aRR=3.05; 95%CI=1.86-5.07), older age(p-trend <0.0001 across age groups; RR=2.32; 95%CI=1.54-3.48 comparing 40-49 to 20-29 years-old), asthma (aRR=1.80; 95%CI=1.29-2.52), diabetes (treated without insulin aRR=2.39; 95%CI=1.46-3.89, requiring insulin aRR=31.89; 95%CI=16.78-60.60), hypertension (aRR=2.02; 95%CI=1.29-3.16), IDU (aRR=3.97; 95%CI=2.44-6.43), and problematic alcohol use (aRR=3.05; 95%CI=1.86-5.02), were factors significantly associated with higher risk of hospital admission.

**Table 3.** Factors associated with hospitalization status in multivariable Poisson regression analysis with robust error variance among women of reproductive age (15-49 years-old), BC COVID-19 Cohort^‡⍰^.

| Variable | Category | 2020.JAN.26–2021.JAN.15<br>(N=17,036; hospitalized=191) |  |
| --- | --- | --- | --- |
|  |  | aRR (95%CI)* | P† |
| **Age<sup>\*\*</sup>**<br>(Reference group:<br>20-29 years) | <20 years | 0.24 (0.06-0.95) | **0.0424** |
|  | 30-39 years | 1.99 (1.35-2.94) | **0.0005** |
|  | 40-49 years | 2.32 (1.54-3.48) | **<0.0001** |
| **Asthma** |  | 1.80 (1.29-2.52) | **0.0005** |
| **Diabetes** (vs. non-diabetic) | Non-insulin | 2.39 (1.46-3.89) | **0.0005** |
|  | Insulin<sup>1</sup> | 31.89 (16.78-60.60) | **<0.0001** |
| **Hypertension** |  | 2.02 (1.29-3.16) | **0.0020** |
| **Injection drug use<sup>2</sup>** |  | 3.97 (2.44-6.43) | **<0.0001** |
| **Pregnancy** |  | 2.69 (1.42-5.07) | **0.0023** |
| **Problematic alcohol use<sup>3</sup>** |  | 3.05 (1.86-5.02) | **<0.0001** |

<sup>‡</sup> Individuals residing in long term care facilities were excluded.
<sup>‡</sup> 657 observations with missing pregnancy status were removed from original sample (N=17,693).
\* Incidence rate ratios adjusted for the variables present in the table.
\*\* P-trend >0.0001 (age groups assessed as pseudo-continuous values)
† Wald's test.
<sup>1</sup> Any type; includes concomitant treatment with antihyperglycemic agents.
<sup>2</sup> Janjua NZ, Islam N, Kuo M, et al. Identifying injection drug use and estimating population size of people who inject drugs using healthcare administrative datasets. *Int J Drug Policy*. 2018;55:31-39.
https://doi.org/10.1016/j.drugpo.2018.02.001.
<sup>3</sup> Janjua NZ, Kuo M, Yu A, Alvarez M, Wong S, Cook D, et al. The Population Level Cascade of Care for Hepatitis C in British Columbia, Canada: The BC Hepatitis Testers Cohort (BC-HTC). *EBioMedicine* 2016;12:189–95.
https://doi.org/10.1016/j.ebiom.2016.08.035.

In the analysis stratified by age (Table 4), diabetes, and problematic alcohol use were associated with hospitalization across all age groups, with higher risk among younger groups that decreased with older age. Male sex, cancer and hypertension were associated with higher risk of hospital admission among those over 40 years of age. In the analysis restricted to hospitalizations lasting more than two days (Supplemental Table S4) did not show different findings compared with the overall analysis.

**Table 4.**
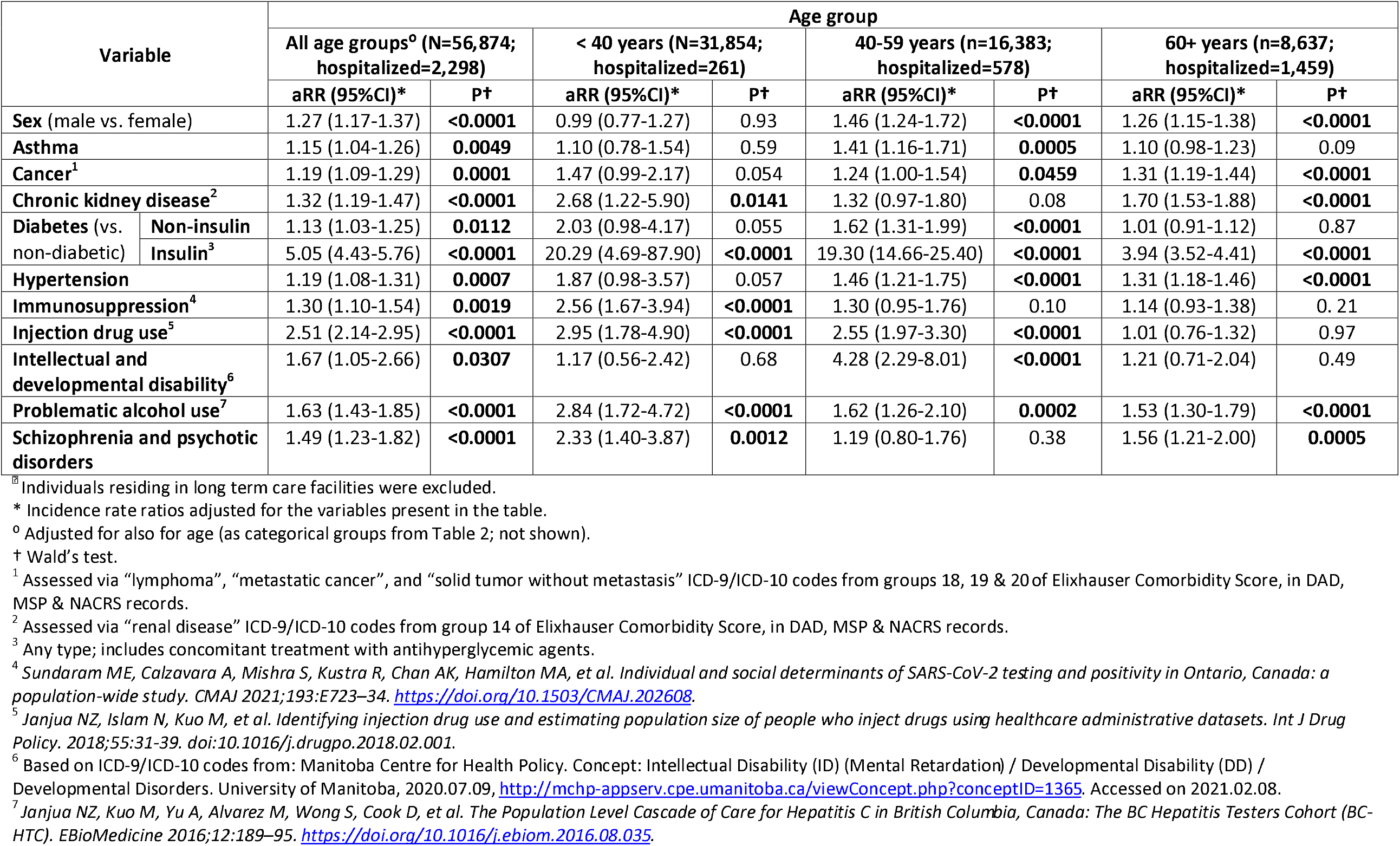
Factors associated with hospitalization status in multivariable Poisson regression analysis with robust error variance among confirmed cases, BC COVID-19 Cohort ^⍰^, stratified by age group.

## Discussion

In this large population-based analysis of all COVID-19 cases in BC, older age, male sex, pregnancy, and numerous chronic co-morbidities and other health-related conditions such as substance use were associated with increased hospitalization risk. Older age was the strongest predictor of hospital admission with risk increasing more than 40 fold for the oldest group compared to the 20-29 years. In addition to well-characterized co-morbidities, we found that injection drug use,^11^ problematic alcohol use, schizophrenia, and psychotic disorders,^12^ as well as intellectual and developmental disability, were independently associated with higher risk of hospitalization, highlighting syndemic of substance use, mental illness and COVID-19. Our findings have important implications for the vaccination program to prevent COVID-19 infection and severe outcomes and subsequently reduce hospital burden and mortality and were used to inform COVID-19 vaccination program in BC.^13^

Our analysis confirms findings from other studies evaluating risk factors for severe COVID-19 outcomes, although ours is one of the few population-based analyses(i.e., includes all COVID-19 diagnoses in a jurisdiction). Most evaluations have also focused on in-hospital mortality, rather than morbidity. In addition to older age and male sex, a wide range of co-morbidities were associated with a higher risk of hospitalization, reflecting similar findings from other studies.^7, 14–19^ These co-morbidities included asthma, chronic kidney disease, diabetes, cancer, immunosuppression and substance use. Associations between most co-morbidities and hospitalization were stronger at younger ages, highlighting the overall low risk of hospitalization among younger people without pre-existing co-morbidities, despite the findings that the hospitalization risk increased with age in the overall population. The highest risk was observed in people of older ages with co-morbidities with incremental risk by age. Several biological studies have identified sex and age differences in biological pathways related to SARS-CoV-2 infection and support our findings. ^20–22^ Pregnancy has been previously identified as a potential risk factor for ICU admission^23,24^ and severe disease,^25^ but most studies have been limited to pregnant women who were already hospitalized (including for non-COVID-19 reasons such as childbirth).^23,26^ This finding could be in part the result of a lower clinical threshold for hospitalization of pregnant patients.

Insulin-dependent diabetes, in particular in the stratum younger than 40 years, was associated with higher risk of hospitalization. To our knowledge this is the first report observing this phenomenon, though insulin use and increased risk of COVID-19-related death was described earlier. Further research is needed to better characterize this finding.

Our analysis also highlights the intersection between the two ongoing public health emergencies in BC: the COVID-19 and the opioid overdose epidemics. The COVID-19 pandemic has exacerbated the pre-existing opioid epidemic, through mechanisms such as disruption of harm reduction services,^27^ with BC experiencing a record high number of illicit drug toxicity deaths in 2020.^28^ Our findings indicate that individuals at high risk of overdose, as indicated by IDU, are also at higher risk of COVID-19 hospitalization. This is the first study investigating impact of COVID-19 on people who inject drugs. IDU was the second co-morbidity most strongly associated with hospitalization in our analysis. Similarly problematic alcohol use, and schizophrenia and psychotic disorders were also associated with higher risk of hospitalization. The higher risk of COVID-19-related hospitalization highlights the syndemic of substance use, mental illness and COVID-19. Many underlying social conditions such as unstable housing, lower socioeconomic status and many co-occurring co-morbidities may have exacerbated the effect of COVID-19 infection among these individuals. Prioritization of vaccination for this population group could reduce disparities and increased risk of hospitalization.

Our analysis had several limitations. We relied on administrative data to identify patient characteristics and co-morbidities; this may have led to some level of misclassification. Similarly, for the same reason, it is not possible to evaluate clinical severity of the event leading to hospital admission. Further, we did not have information on socioeconomic status, race/ethnicity, and obesity. Also, given that the evaluation of the COVID-19 status depends on the BC diagnostic testing guidelines (varying over time to focus on symptom-based assessment since April 21st, 2020),^29^ selective ascertainment of symptomatic cases is expected, resulting in exclusion of asymptomatic cases.^30^

In conclusion, older age, male sex, pregnancy, and various comorbidities and health-conditions including substance use were associated with higher risk of hospital admission in this population-based analysis. These findings have informed the COVID-19 vaccination program rollout in BC and will be useful for informing the prioritization of vaccination in other jurisdictions to prevent infection and severe outcomes.^13^ In addition, these findings will also inform monitoring of individual patients by their healthcare providers at higher risk of severe outcomes. Finally there is a need for further characterizing syndemics of substance use, mental illness and COVID-19.

## Data Availability

The study is based on data contained in various provincial registries and databases. Access to data could be requested through the BC Centre for Disease Control Institutional Data Access for researchers who meet the criteria for access to confidential data. Requests for the data may be sent to.

## Contributors

HAVG and NZJ contributed to study conception, design, and analyzed data. NZJ, NP and MK were involved in funding acquisition. NJ, SD, EG, JW, MC, and MK were involved in data acquisition. All authors contributed to data interpretation. HAVG was responsible for the first draft of the manuscript. All authors contributed to critical revision of the manuscript. All authors had access to and verified all the data and accept responsibility for the decision to submit for publication. The database for the study has been accessed and verified by HAVG and NZJ. All authors had full access to all the data and had final responsibility for the decision to submit for publication.

## Disclaimer

All inferences, opinions, and conclusions drawn in this report are those of the authors, and do not reflect the opinions or policies of the Data Steward(s).

## Acknowledgements

We acknowledge the assistance of the Provincial Health Services Authority, BC Centre for Disease Control, BC Ministry of Health and Regional Health Authority staff involved in data access, procurement, and management. We gratefully acknowledge the residents of British Columbia whose data are integrated in the British Columbia COVID-19 Cohort (BCC19C).

