## Supplementary data for "Mental health and substance use associated with hospitalization among people with laboratory confirmed diagnosis of COVID-19 in British Columbia: a population-based cohort study"

**Appendix A. BC COVID-19 Cohort** (BCC19C)

The BC COVID-19 Cohort (BCC19C) is a surveillance platform that integrates COVID-19 data with administrative health datasets (Table S1). The platform was established in 2020 in order to support the public health response to the COVID-19 pandemic. The BCC19C is a dynamic cohort in which COVID-19 datasets are updated daily and most administrative health datasets are updated weekly/monthly. The majority of the integrated datasets are population-based (cover all individuals in the province). Datasets are de-identified and linked together through a combination of probabilistic and deterministic matching algorithms, as described previously.^[1]^

The BCC19C is a collaboration between the Provincial Health Services Authority (PHSA), BC Centre for Disease Control (BCCDC), and the BC Ministry of health (MOH). The platform was established under BCCCDC’s public health mandate. This study was also reviewed and approved by the Behavioral Research Ethics Board at the University of British Columbia (H20-02097).

**Table S1 -** **Data Sources integrated within the BC COVID-19 Cohort (BCC19C)**

| **British Columbia Centre for Disease Control (BCCDC), Provincial Heath Services Authority (PHSA) and Regional Health Authority data sources:** | **Data Date Ranges:** |
| --- | --- |
| Integrated COVID-19 laboratory dataset (SARS-CoV2 tests from private/public labs) ^S1^ | Jan,2020-onward |
| COVID-19 surveillance case data (information collected on all probable/confirmed cases as part of public health follow up) ^S2^ | Jan,2020-onward |
| Provincial COVID-19 Monitoring Solution (critical and non-critical care hospital census data) ^S3^ | Jan,2020-onward |
| Provincial Immunizations Registry (COVID-19 vaccination data) ^S4^ | Dec,2020-onward |
| Provincial Laboratory Information Solution (laboratory tests from private/public labs) ^S5^ | Jan,2020-onward |
| Public Health Reporting Data warehouse (Influenza laboratory tests)^S6^ | Jan,2008-onward |
| Emergency department visits (hospital-based and community-based ambulatory care) | Mar,2020-onward |
| Ministry of Health (MoH) Administrative Data Sources: | **Data Date Ranges:** |
| Client Roster (CR) (registry of enrollment in the universal public health insurance plan including residential history) ^S7^ | 2008/9-onward |
| Discharge Abstracts Database (DAD) (hospital discharge records) ^S8^ | 2008/9-onward |
| Medical Services Plan (MSP) (physician diagnostic and billing data for services provided through universal public health insurance plan) ^S9^ | 2008/9-onward |
| PharmaNet (Pharma) (prescription drugs dispensed from community pharmacies, includes medications covered by public and private insurance plans) ^S10^ | 2008/9-onward |
| BC Vital Statistics (VS) (deaths registry) ^S11^ | 2008/9-onward |
| National Ambulatory Care Reporting System (NACRS) (hospital-based and community-based ambulatory care) ^S12^ | 2011/12–onward |
| Chronic Disease Registry ^S13^ | 2008/9-2018/19 |
| 811 Calls (respiratory calls only) ^S14^ | 2014-onward |
| Health System Matrix ^S15^ | 2018/19-onward |
| Population Grouper Methodology ^S16^ | 2008/9-onward |

**References (Data Sources)**

S1. British Columbia Centre for Disease Control [creator]. Integrated COVID-19 laboratory dataset (SARS-CoV2 tests from private/public labs). Public Health Reporting Data Warehouse, British Columbia Centre for Disease Control [publisher] (2020). 2021.
S2. British Columbia Centre for Disease Control [creator]. COVID-19 surveillance case data. British Columbia Centre for Disease Control [publisher]. (2020). 2021.
S3. Provincial Health Services Authority [creator]. Provincial COVID-19 Monitoring Solution. Provincial Health Services Authority [publisher]. (2020). 2021.
S4. Provincial Health Services Authority [creator]. Provincial Public Health Information Systems [publisher]. (2020). 2021.
S5. Provincial Health Services Authority [creator]. COVID-19 vaccination data. Provincial Immunizations Registry, Provincial Public Health Information Systems [publisher]. (2020). 2021.
S6. British Columbia Centre for Disease Control [creator]. Respiratory datamart, Public Health Reporting Data Warehouse, British Columbia Centre for Disease Control [publisher] (2020). 2021.
S7. British Columbia Ministry of Health [creator]. Client Roster (Client Registry System/Enterprise Master Patient Index). British Columbia Ministry of Health [publisher]. Data Extract. MOH (2020). 2021. <https://www2.gov.bc.ca/gov/content/health/health-forms/online-services>
S8. British Columbia Ministry of Health [creator]. Discharge Abstract Database (Hospital Separations). British Columbia Ministry of Health [publisher]. Data Extract. MOH (2020). 2021. <https://www2.gov.bc.ca/gov/content/health/health-forms/online-services>
S9. British Columbia Ministry of Health [creator]. Medical Services Plan (MSP) Payment Information File. British Columbia Ministry of Health [publisher]. Data Extract. MOH (2020). 2021.<https://www2.gov.bc.ca/gov/content/health/health-forms/online-services>
S10. British Columbia Ministry of Health [creator]. PharmaNet. British Columbia Ministry of Health [publisher]. Data Extract. MOH (2020). 2021. <https://www2.gov.bc.ca/gov/content/health/health-forms/online-services>
S11. BC Vital Statistics Agency [creator]. Vital Statistics Deaths. BC Vital Statistics Agency [publisher]. Data Extract. BC Vital Statistics Agency (2020). 2021. <https://www2.gov.bc.ca/gov/content/health/health-forms/online-services>
S12. British Columbia Ministry of Health [creator]. National Ambulatory Care Reporting System. British Columbia Ministry of Health [publisher]. Data Extract. MOH (2020). 2021.

<https://www2.gov.bc.ca/gov/content/health/health-forms/online-services>

S13. British Columbia Ministry of Health [creator]. Chronic Disease Registry. British Columbia Ministry of Health [publisher]. Data Extract. MOH (2020). 2020.

<https://www2.gov.bc.ca/gov/content/health/health-forms/online-services>

S14. British Columbia Ministry of Health [creator]. 811 calls. British Columbia Ministry of Health [publisher]. Data Extract. MOH (2020). 2021.

<https://www2.gov.bc.ca/gov/content/health/health-forms/online-services>

S15. British Columbia Ministry of Health [creator]. Health System Matrix. British Columbia Ministry of Health [publisher]. Data Extract. MOH (2020). 2021.

<https://www2.gov.bc.ca/gov/content/health/health-forms/online-services>

S16. British Columbia Ministry of Health [creator]. Population Grouper Methodology. British Columbia Ministry of Health [publisher]. Data Extract. MOH (2020). 2021.

<https://www2.gov.bc.ca/gov/content/health/health-forms/online-services>

**Appendix B. Obstetric codes (liveborn and stillborn)**

Based on “ICD-9 and ICD-10 Codes for Liveborn and Stillborn (Baby)”, from *Manitoba Child Health Atlas Update (2008)*^[2]^; revised codes are as follows:

- DAD1 ICD-9-CM diagnostic codes starting with: V3, V27; exact code 7799
- DAD2 ICD-10-CA diagnostic codes starting with: Z37; exact code P95

**Appendix C. Definitions for comorbidity variables derived from administrative datasets**

**Administrative data sources:**

A) Chronic disease registry (CDR)^5^ – Available information includes diagnosis date(s)
B) Medical Services Plan (MSP)^1^ – ICD-9 billing/diagnostic codes.
C) Discharge Abstract Database (DAD)^3^ – DAD1 contains ICD-9 coded hospitalization data and DAD2 contains ICD-10 coded hospitalization data.
D) National Ambulatory Care Reporting System (NACRS)^4^ – Contains ICD-10 coded diagnostic codes.
E) PharmaNet^2^ – Each medication is identified with a drug identification number (DINPIN).

**All comorbidities were identified according to the information extracted from CDR, with the following exceptions:**

**1. Cirrhosis**

- MSP ICD-9 diagnostic codes: starting with 4562; exact codes 4560, 4561, 56723, 5722 , 5724, 5712, 5728, 5715, 7895, 07044, 5713
- DAD1 ICD-9-CM diagnostic codes: starting with 4562; exact codes 4560, 4561, 56723, 5722, 5723, 5724, 5712, 5728, 5715, 7895, 07044, 5713
- DAD2/NACRS ICD-10-CA diagnostic codes: starting with K703; exact codes: I850 ,K652, K721, K729, K766, K767, K7460, K7469, R18, I98.20, I98.3, K704

**2. Cancer (ever)**

Any occurrence of groups 18, 19, or 20 (“lymphoma”, “metastatic cancer”, and “solid tumor without metastasis”) of Elixhauser comorbidity index^[3]^.

**3. Chronic kidney disease**

As per group 14 (“renal failure”) of Elixhauser comorbidity index^8^.

**4. Diabetes (DM)**

Diabetes status as per CDR; classified as:

- DM requiring insulin: PharmaNet DINPINs 1934074, 1959220, 2024284, 586714, 2024233, 446564, 612227, 1934112, 612197, 632651, 632686, 1986085, 1986805, 1986813, 2025256, 1959239, 2024268, 587737, 2024225, 446572, 612235, 1934066, 646148, 2024241, 446580, 612278, 1934090, 2024403, 2241310, 2415089, 2403447, 795879, 2024217, 1959212, 2025248, 889121, 2024446, 1962639, 2024292, 1962655, 2024306, 1962663, 2024322, 1962647, 2024314, 889091, 889105, 889113, 2466864, 644358, 733075, 2024276, 513644, 2275872, 514551, 552275, 612170, 1985949, 2022249, 2275864, 514535, 612162, 612359, 2244353, 2245397, 2377209, 2460416, 2460424, 2460408, 2265435, 2265443, 2467879, 2467887, 2474875, 2412829, 2271842, 2245689, 2276410, 2251930, 2444844, 2294338, 2444852, 2461528, 2441829, 2493373, 2478293, 2279479, 2279460, 2279487, 2294346, 2229704, 2469901, 2229705, 2469898, 2233562, 2241283, 2403412, 2469871, 2439611, 2470152, 2240294, 2240295, 2403420, 2240297, 2403439, 632694, 650935, 1986821, 2022230, 552259, 614416, 1985957, 1985981, 612189, 552267, 628301, 723789, 1985930, 446610, 612219, 612200, 446599, 612243, 1934104, 446602, 612251, 1934082, 773654, 1985973, 632678, 1985965, 5894, 6009, 274119, 274127, 275409, 275417, 275425, 539201, 539244, 542911, 542938, 542946, 546348, 554820, 648094, 999717, 1986791, 45230001, 45230002, 45230003, 45230004, 45230005, 45230006, 45230007, 45230008, 45230009, 45230010, 45230011, 45230012, 45230013, 47450001, 47450002, 47450003, 47450004, 47450005, 47450006, 47450007, 66123203, 66124134, 66124135, 66124225, 66124582, 45230014, 45230015, 45230016, 46340034, 46340035, 46340036, 46340037, 47450008, 47450009.
- DM treated without insulin: Any record lacking the aforementioned DINPINs.

**5. Immunosuppression**

Based on Sundaram et al. 2020^[4]^. Includes any occurrence in DAD/NACRS of:

- Antibody- or leukocyte-related immune disorders: ICD-10: D80 - D84, D89.8, D89.9
- Neutropenia - ICD-10: D70
- Abnormality of leukocytes - ICD-10: D71, D72
- Splenomegaly - ICD-10: D73.0 - D73.2
- Asplenia - ICD-10: Q890
- Sickle cell syndrome - ICD-10: D570 - D572
- Transplant: Organ or tissue replaced by transplant - ICD-9 (MSP): V42; Transplanted organ and tissue status - ICD-10: Z94
- AIDS/HIV – As per group 17 (“AIDS/HIV”) of Elixhauser comorbidity index^8^.

**6. Injection Drug Use^[5]^**

Diagnosis age between 11 and 65:

- MSP ICD-9 diagnostic codes: starting with 292, 970, 3040, 3041, 3042, 3044, 3045, 3046, 3047, 3048, 3049, 3054, 3055, 3056, 3057, 3059, 6483, 7960, 9621, 9650, 9658, 9663, 9664, 9670, 9684, 9685, 9694, 9696, 9697, 9698, 9699, E8500, V6542; exact code V6542
- DAD1 ICD-9-CM diagnostic codes: starting with 292, 970, 3040, 3041, 3042, 3044, 3045, 3046, 3047, 3048, 3049, 3054, 3055, 3056, 3057, 3059, 6483, 7960, 9621, 9650, 9658, 9663, 9664, 9670, 9684, 9685, 9694, 9696, 9697, 9698, 9699, E8500, V6542; exact code V6542
- DAD2/NACRS ICD- 10-CA diagnostic codes: starting with F11, F13, F14, F15, F19, R781, R782, T387, T400, T401, T402, T403, T404, T405, T406, T408, T409, T412, T423, T424, T425, T426, T427, T428, T436, T438, T439, T507; exact codes: R781, R782, T387, T400, T401, T402, T403, T404, T405, T406, T408, T409, T412, T423, T424, T425, T426, T427, T428, T436, T438, T439, T507
- NACRS complaint codes: starting with 751, 753

**7. Problematic Alcohol Use^[6]^**

- MSP ICD-9 diagnostic codes: starting with 291, 303, 3050, 3575, 4255
- DAD1 ICD-9-CM: starting with 291, 303, 3050,3575, 4255
- DAD2/NACRS ICD-10-CA: starting with F10, E244, G312, G621, G721, I426, Z502, Z714.

**8. Schizophrenia and psychotic disorders^[7]^**

2 MSP occurrences, or 1 DAD, or 1 NACRS

- MSP ICD-9 diagnostic codes: starting with 295, 2988, 2989, 2970, 2972, 2971, 2973, 2978, 2979
- DAD1 ICD-9-CM: starting with 295, 2988, 2989, 2970, 2972, 2971, 2973, 2978, 2979
- DAD2/NACRS ICD-10-CA: starting with F20, F21, F22, F23, F24, F25, F28, F29.

**Appendix D. Analytic sample flow diagram**

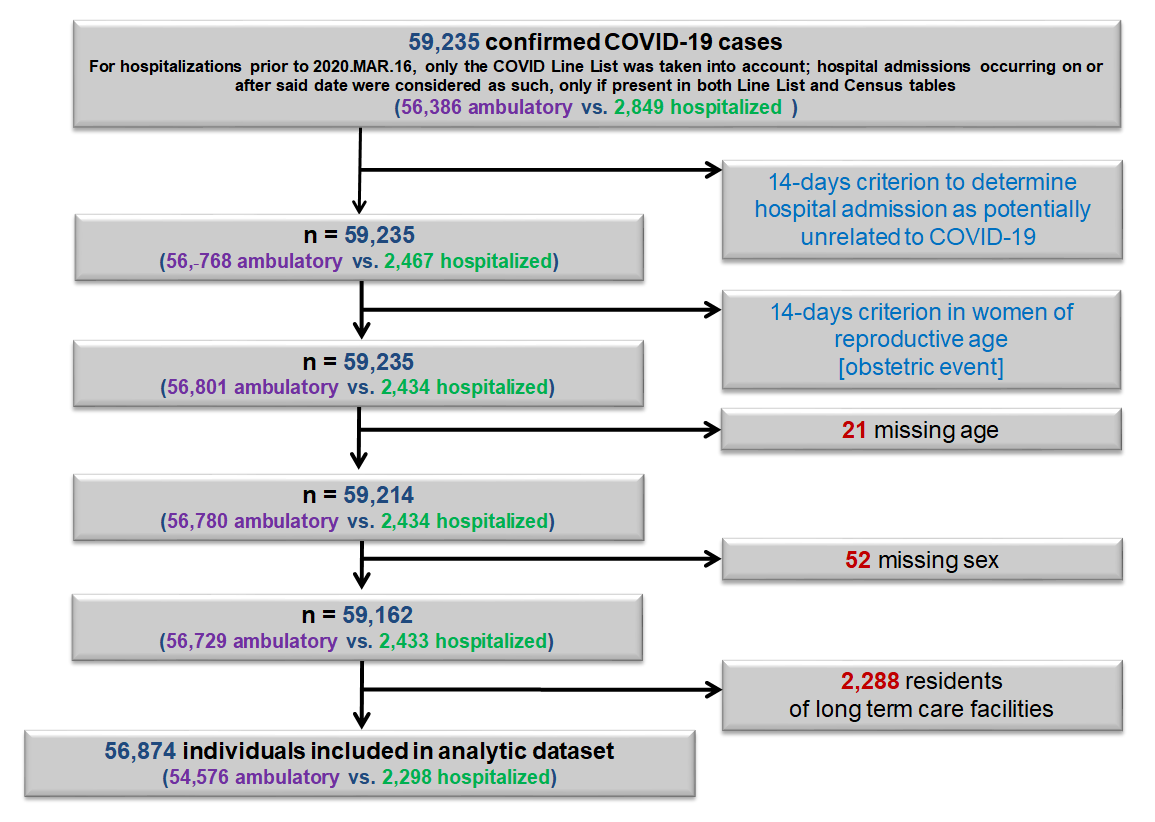

**Table S2 - Factors associated with hospitalization status in multivariable Poisson regression analysis with robust error variance among confirmed cases, BC COVID-19 Cohort^⌂^, stratified by time period**

| **Variable** | **Category** | | **2020.JAN.26–2021.JAN.15 (N=56,874; hospitalized=2,298)** | | **2020.JAN.26–2020.AUG.01 (N=3,310; hospitalized=410)** | | **2020.AUG.02–2021.JAN.15 (N=53,564; hospitalized=1,888)** | |
| --- | --- | --- | --- | --- | --- | --- | --- | --- |
|  |  |  | **aIRR (95%CI)*** | **P†** | **aIRR (95%CI)*** | **P†** | **aIRR (95%CI)*** | **P†** |
| **Sex** (vs. female) | Male | | 1.27 (1.17-1.37) | **<0.0001** | 1.34 (1.14-1.59) | **0.0006** | 1.24 (1.14-1.35) | **<0.0001** |
| **Age**^‡^  (Reference group:  20-29 years) | <20 years | | 0.51 (0.30-0.85) | **0.0103** | *NE* | - | 0.60 (0.35-1.01) | 0.0534 |
|  | 30-39 years | | 3.06 (2.32-4.03) | **<0.0001** | 2.46 (1.21-5.01) | **0.0130** | 3.03 (2.25-4.09) | **<0.0001** |
|  | 40-49 years | | 4.45 (3.40-5.82) | **<0.0001** | 4.05 (2.04-8.03) | **<0.0001** | 4.31 (3.22-5.77) | **<0.0001** |
|  | 50-59 years | | 8.05 (6.22-10.41) | **<0.0001** | 5.86 (3.03-11.37) | **<0.0001** | 7.85 (5.94-10.37) | **<0.0001** |
|  | 60-69 years | | 14.98 (11.58-19.37) | **<0.0001** | 11.35 (5.88-21.91) | **<0.0001** | 14.48 (10.96-19.14) | **<0.0001** |
|  | 70-79 years | | 28.15 (21.64-36.61) | **<0.0001** | 18.72 (9.63-36.36) | **<0.0001** | 27.51 (20.67-36.61) | **<0.0001** |
|  | 80+ years | | 43.68 (33.41-57.10) | **<0.0001** | 27.64 (13.98-54.62) | **<0.0001** | 43.09 (32.21-57.64) | **<0.0001** |
| **Asthma** | | | 1.15 (1.04-1.26) | **0.0049** | 1.27 (1.04-1.55) | **0.0182** | 1.13 (1.02-1.26) | **0.0236** |
| **Cancer^1^** | | | 1.19 (1.09-1.29) | **0.0001** | 1.10 (0.91-1.33) | 0.31 | 1.19 (1.08-1.32) | **0.0005** |
| **Chronic kidney disease^2^** | | | 1.32 (1.19-1.47) | **<0.0001** | 1.15 (0.93-1.41) | 0.20 | 1.31 (1.17-1.48) | **<0.0001** |
| **Diabetes** (vs. non-diabetic) | | Non-insulin | 1.13 (1.03-1.25) | **0.0112** | 1.12 (0.91-1.37) | 0.29 | 1.15 (1.04-1.29) | **0.0092** |
|  |  | Insulin**^3^** | 5.05 (4.43-5.76) | **<0.0001** | 3.11 (2.34-4.14) | **<0.0001** | 5.61 (4.85-6.49) | **<0.0001** |
| **Hypertension** | | | 1.19 (1.08-1.31) | **0.0007** | 1.09 (0.89-1.34) | 0.40 | 1.22 (1.09-1.36) | **0.0005** |
| **Immunosuppression^4^** | | | 1.30 (1.10-1.54) | **0.0019** | 1.14 (0.81-1.60) | 0.46 | 1.33 (1.10-1.59) | **0.0028** |
| **Injection drug use^5^** | | | 2.51 (2.14-2.95) | **<0.0001** | 2.46 (1.63-3.70) | **<0.0001** | 2.63 (2.21-3.14) | **<0.0001** |
| **Intellectual & developmental disability^6^** | | | 1.67 (1.05-2.66) | **0.0307** | 2.10 (1.17-3.77) | **0.0130** | 1.50 (0.87-2.58) | **0.14** |
| **Problematic alcohol use^7^** | | | 1.63 (1.43-1.85) | **<0.0001** | 1.57 (1.23-2.02) | **0.0003** | 1.65 (1.43-1.91) | **<0.0001** |
| **Schizophrenia and psychotic disorders** | | | 1.49 (1.23-1.82) | **<0.0001** | 0.70 (0.42-1.14) | 0.15 | 1.67 (1.35-2.06) | **<0.0001** |

**^⌂^** Individuals residing in long term care facilities were excluded.

* Incidence rate ratios adjusted for the variables present in the table.

† Wald’s test.

^‡^ P-trend >0.0001 (age groups assessed as pseudo-continuous values)

^1^ Assessed via “lymphoma”, “metastatic cancer”, and “solid tumor without metastasis” ICD-9/ICD-10 codes from groups 18, 19 & 20 of Elixhauser Comorbidity Score, in DAD, MSP & NACRS records.

^2^ Assessed via “renal disease” ICD-9/ICD-10 codes from group 14 of Elixhauser Comorbidity Score, in DAD, MSP & NACRS records.

^3^ Any type; includes concomitant treatment with antihyperglycemic agents.

^4^ *Sundaram ME, Calzavara A, Mishra S, Kustra R, Chan AK, Hamilton MA, et al. Individual and social determinants of SARS-CoV-2 testing and positivity in Ontario, Canada: a population-wide study. CMAJ 2021;193:E723–34.* [*https://doi.org/10.1503/CMAJ.202608*](https://doi.org/10.1503/CMAJ.202608)*.*

^5^ *Janjua NZ, Islam N, Kuo M, et al. Identifying injection drug use and estimating population size of people who inject drugs using healthcare administrative datasets. Int J Drug Policy. 2018;55:31-39.* [*https://doi:10.1016/j.drugpo.2018.02.001*](https://doi:10.1016/j.drugpo.2018.02.001).

^6^ Based on ICD-9/ICD-10 codes from: Manitoba Centre for Health Policy. Concept: Intellectual Disability (ID) (Mental Retardation) / Developmental Disability (DD) / Developmental Disorders. University of Manitoba, 2020.07.09, <http://mchp-appserv.cpe.umanitoba.ca/viewConcept.php?conceptID=1365>. Accessed on 2021.02.08.

^7^ *Janjua NZ, Kuo M, Yu A, Alvarez M, Wong S, Cook D, et al. The Population Level Cascade of Care for Hepatitis C in British Columbia, Canada: The BC Hepatitis Testers Cohort (BC-HTC). EBioMedicine 2016;12:189–95.* [*https://doi.org/10.1016/j.ebiom.2016.08.035*](https://doi.org/10.1016/j.ebiom.2016.08.035)*.*

*NE:* Not examined due lack of observations presenting the outcome of interest within the variable’s stratum.

**Table S3 - Factors associated with hospitalization status in multivariable Poisson regression analysis with robust error variance among women of reproductive age (15-49 years-old), BC COVID-19 Cohort^‡⌂^, stratified by time period**

| **Variable** | **Category** | | **2020.JAN.26–2021.JAN.15 (N=17,036; hospitalized=191)** | | **2020.JAN.26–2020.AUG.01 (N=906; hospitalized=33)** | | **2020.AUG.02–2021.JAN.12 (N=16,130; hospitalized=158)** | |
| --- | --- | --- | --- | --- | --- | --- | --- | --- |
|  |  |  | **aIRR (95%CI)*** | **P†** | **aIRR (95%CI)*** | **P†** | **aIRR (95%CI)*** | **P†** |
| **Age**^**^  (Reference group:  20-29 years) | <20 years | | 0.24 (0.06-0.95) | **0.0424** | *NE* | - | 0.27 (0.07-1.09) | 0.07 |
|  | 30-39 years | | 1.99 (1.35-2.94) | **0.0005** | 1.83 (0.68-5.17) | 0.25 | 1.95 (1.28-2.96) | **0.0019** |
|  | 40-49 years | | 2.32 (1.54-3.48) | **<0.0001** | 3.14 (1.05-9.42) | **0.0408** | 2.10 (1.35-3.26) | **0.0010** |
| **Asthma** | | | 1.80 (1.29-2.52) | **0.0005** | 1.55 (0.68-3.52) | 0.29 | 1.88 (1.31-2.71) | **0.0007** |
| **Diabetes** (vs. non-diabetic) | | Non-insulin | 2.39 (1.46-3.89) | **0.0005** | 2.86 (1.16-6.99) | **0.0216** | 2.17 (1.23-3.85) | **0.0078** |
|  |  | Insulin**^1^** | 31.89 (16.78-60.60) | **<0.0001** | 8.26 (2.40-28.46) | **0.0008** | 34.13 (15.65-74.43) | **<0.0001** |
| **Hypertension** | | | 2.02 (1.29-3.16) | **0.0020** | 1.33 (0.39-4.49) | 0.64 | 2.27 (1.39-3.71) | **0.0011** |
| **Injection drug use^2^** | | | 3.97 (2.44-6.43) | **<0.0001** | 4.82 (1.38-16.80) | **0.0135** | 3.91 (2.28-6.71) | **<0.0001** |
| **Pregnancy** | | | 2.69 (1.42-5.07) | **0.0023** | 2.65 (0.64-10.89) | 0.18 | 2.60 (1.28-5.29) | **0.0083** |
| **Problematic alcohol use^3^** | | | 3.05 (1.86-5.02) | **<0.0001** | 1.82 (0.47-7.07) | 0.38 | 3.30 (1.91-5.71) | **<0.0001** |

**^⌂^** Individuals residing in long term care facilities were excluded.

^‡^ 657 observations with missing pregnancy status were removed from original sample (N=17,693).

* Incidence rate ratios adjusted for the variables present in the table.

^**^ P-trend >0.0001 (age groups assessed as pseudo-continuous values)

† Wald’s test.

^1^ Any type; includes concomitant treatment with antihyperglycemic agents.

^2^ *Janjua NZ, Islam N, Kuo M, et al. Identifying injection drug use and estimating population size of people who inject drugs using healthcare administrative datasets. Int J Drug Policy. 2018;55:31-39.* [*https://doi:10.1016/j.drugpo.2018.02.001*](https://doi:10.1016/j.drugpo.2018.02.001).

^3^ *Janjua NZ, Kuo M, Yu A, Alvarez M, Wong S, Cook D, et al. The Population Level Cascade of Care for Hepatitis C in British Columbia, Canada: The BC Hepatitis Testers Cohort (BC-HTC). EBioMedicine 2016;12:189–95.* [*https://doi.org/10.1016/j.ebiom.2016.08.035*](https://doi.org/10.1016/j.ebiom.2016.08.035)*.*

*NE:* Not examined due lack of observations presenting the outcome of interest within the variable’s stratum.

**Table S4 - Factors associated with hospitalization status in multivariable Poisson regression analysis with robust error variance among confirmed cases, BC COVID-19 Cohort^⌂^, restricted to hospital admissions lasting more than two days**

| **Variable** | **Category** | | **2020.JAN.26–2021.JAN.15**  **(N=56,482; hospitalized=1,906)** | |
| --- | --- | --- | --- | --- |
|  |  |  | **aRR (95%CI)*** | **P†** |
| **Sex** (vs. female) | Male | | 1.30 (1.19-1.41) | **<0.0001** |
| **Age**^‡^  (Reference group:  20-29 years) | <20 years | | 0.20 (0.08-0.49) | **0.0005** |
|  | 30-39 years | | 3.19 (2.30-4.43) | **<0.0001** |
|  | 40-49 years | | 5.20 (3.79-7.12) | **<0.0001** |
|  | 50-59 years | | 9.81 (7.25-13.28) | **<0.0001** |
|  | 60-69 years | | 18.16 (13.43-24.57) | **<0.0001** |
|  | 70-79 years | | 35.23 (25.88-47.94) | **<0.0001** |
|  | 80+ years | | 54.77 (40.01-74.98) | **<0.0001** |
| **Asthma** | | | 1.13 (1.02-1.25) | **0.0251** |
| **Cancer^1^** | | | 1.16 (1.05-1.28) | **0.0027** |
| **Chronic kidney disease^2^** | | | 1.38 (1.23-1.55) | **<0.0001** |
| **Diabetes** (vs. non-diabetic) | | Non-insulin | 1.10 (0.99-1.22) | 0.08 |
|  |  | Insulin**^3^** | 5.31 (4.60-6.12) | **<0.0001** |
| **Hypertension** | | | 1.18 (1.06-1.32) | **0.0029** |
| **Immunosuppression^4^** | | | 1.30 (1.08-1.56) | **0.0056** |
| **Injection drug use^5^** | | | 2.33 (1.95-2.80) | **<0.0001** |
| **Intellectual & developmental disability^6^** | | | 1.96 (1.22-3.15) | **0.0053** |
| **Problematic alcohol use^7^** | | | 1.64 (1.42-1.90) | **<0.0001** |
| **Schizophrenia & psychotic disorders** | | | 1.42 (1.13-1.78) | **0.0023** |

**^⌂^** Individuals residing in long term care facilities were excluded.

* Incidence rate ratios adjusted for the variables present in the table.

† Wald’s test.

^‡^ P-trend >0.0001 (age groups assessed as pseudo-continuous values)

^1^ Assessed via “lymphoma”, “metastatic cancer”, and “solid tumor without metastasis” ICD-9/ICD-10 codes from groups 18, 19 & 20 of Elixhauser Comorbidity Score, in DAD, MSP & NACRS records.

^2^ Assessed via “renal disease” ICD-9/ICD-10 codes from group 14 of Elixhauser Comorbidity Score, in DAD, MSP & NACRS records.

^3^ Any type; includes concomitant treatment with antihyperglycemic agents.

^4^ *Sundaram ME, Calzavara A, Mishra S, Kustra R, Chan AK, Hamilton MA, et al. Individual and social determinants of SARS-CoV-2 testing and positivity in Ontario, Canada: a population-wide study. CMAJ 2021;193:E723–34.* [*https://doi.org/10.1503/CMAJ.202608*](https://doi.org/10.1503/CMAJ.202608)*.*

^5^ *Janjua NZ, Islam N, Kuo M, et al. Identifying injection drug use and estimating population size of people who inject drugs using healthcare administrative datasets. Int J Drug Policy. 2018;55:31-39. doi:10.1016/j.drugpo.2018.02.001*.

^6^ Based on ICD-9/ICD-10 codes from: Manitoba Centre for Health Policy. Concept: Intellectual Disability (ID) (Mental Retardation) / Developmental Disability (DD) / Developmental Disorders. University of Manitoba, 2020.07.09, <http://mchp-appserv.cpe.umanitoba.ca/viewConcept.php?conceptID=1365>. Accessed on 2021.02.08.

^7^ *Janjua NZ, Kuo M, Yu A, Alvarez M, Wong S, Cook D, et al. The Population Level Cascade of Care for Hepatitis C in British Columbia, Canada: The BC Hepatitis Testers Cohort (BC-HTC). EBioMedicine 2016;12:189–95.* [*https://doi.org/10.1016/j.ebiom.2016.08.035*](https://doi.org/10.1016/j.ebiom.2016.08.035)*.*
